# Unraveling diagnostic co-morbidity makeup of each HF category as characteristically derived by ECG- and ECHO-findings

**DOI:** 10.1101/2021.09.30.21264236

**Authors:** Azfar Zaman, Marta Afonso Nogueira, Erzsebet Szabo, Aniko Berta-Szabo, Giuseppe Biondi Zoccai, Niall Campbell, Georgios Koulaouzidis, Dionissios Tsipas, Istvan Kecskes

## Abstract

**Background:** Echocardiography (ECHO) is not widely available in primary care, the key structural (chamber enlargements) and functional abnormality are not easily available precluding the ability to diagnose HF other than through mainly symptomatic means. The opportunity for earlier detection of HF is lost.

**Methods:** Using a unique database, the etiology of HF is explored by prevalence analysis to unravel the diagnostic makeup of each HF category. Various relationships and patterns of comorbidities have been extracted between the Electrocardiogram (ECG) and ECHO parameters that contribute to HF, those relationships are then confirmed and categorized by a Principal Component Analysis (PCA). Finally, it was summarized what type of non-invasive ECG-like device should be used in primary care to better diagnose HF.

**Results:** The sensitivity of abnormal ECHO reaches 92% over the abnormal ECG of 81% in the detection of HF. The first five PCA are discovered, which cover 49% of all the variance. Left atrial enlargement is the most representative finding in the overall comorbidity rate, which coincides with the probability direction of HF (3^rd^ as input, 1^st^ as finding in the coefficients), and reaches the highest (250%) prevalence increase in function of decreasing LVEF.

**Conclusions:** The core structural and functional abnormalities diagnosed by ECHO with the ECG interpretation provide sufficient information to diagnose “consider HF” in primary care. This paper overview of a novel bio-signal-based system supported by Artificial Intelligence, able to replicate Echo-findings, predict HF and indicates its phenotype, suitable for use in Primary Care.

## 1 Introduction

Heart Failure (HF) is a heterogeneous syndrome with various etiologic and pathophysiologic factors that define the three main categories, primarily supported by Echocardiogram (ECHO) to determine Left Ventricle Ejection Fraction (LVEF) and to a lesser degree by Electrocardiogram (ECG) as found in guidance and research articles. ^1,2,3,4,5,6,7,8,9^ Failure to understand these abnormalities can undermine detection leading to a failure or delay of diagnosis, and misdirected treatment.

According to recent European Society of Cardiology (ESC) diagnostic guidelines,^1^ the categorization of HF relies primarily on LVEF: 1) reduced, HFrEF (LVEF ≤40%); 2) mildly-reduced, HFmrEF (LVEF between 41% to 49%); and 3) preserved, HFpEF (LVEF of ≥50%). However, LVEF as a measure of HF is limited, as LVEF alone it does not explain the underlying disease characteristics and conditions, for example, LVEF estimates global function but does not indicate Left Ventricular (LV) volume or stroke volume (SV).^4,10^ Also, LVEF is not sufficiently sensitive for subtle LV systolic dysfunction (LVSD) caused by regional Wall Motion Abnormality (WMA) present in HFpEF, better detected by mitral annular systolic excursion or systolic velocities or LV global longitudinal strain (GLS). ^10,11,12^.

Rather, LVEF has to be obtained with an assessment of LVSD and Dilated Cardiomyopathy (DCM) supported by many other structural and functional abnormality and pathophysiological characteristics, such as Left Atrial Enlargement (LAE), Ventricular Hypertrophy (LVH), and Diastolic Dysfunction (DD).^1,2,3,4,13^ In addition, the phenotype of functional abnormalities is strongly connected to Atrial Fibrillation (AFib),^7,8,9,14^ coronary artery disease (CAD), WMA, Aortic Stenosis (AS), and other valve diseases, e.g., Mitral Regurgitation (MR),^15^ as well as Pulmonary Hypertension (PH).^16,17^ The measurement of these findings including LVEF are primarily obtained from Echocardiography.

Not surprisingly, guidelines ^1,3,4^ make little reference for use of ECG which has the low diagnostic capability for HF, primarily due to its poor sensitivity for heart structural abnormalities,^18,19^ especially for left atrial enlargement,^20,21^ and poor sensitivity or specificity for heart functional abnormalities.^22,23,24,25^ Alone, ECG’s inherent diagnostic limitations for atrial and ventricular enlargements make it unsuitable for the detection of HF in primary care settings.

Lacking a suitable device able to detect structural and functional abnormalities required to confirm HF and its categories that are also indicated for use in Primary Care. The current understanding and prediction of HF at the Primary Care level is typically limited to more protracted symptomatic-based assessments leading to a clinical prognosis. Any opportunity for the earlier detection of HF is lost.

The objective of this study is to discover what type of diseases/abnormalities should be detected from bio-signals to support HF diagnosis in Primary Care without ECHO. This study investigate the key abnormalities that enable HF classification. First, we explore the etiology of each HF phenotype, then investigate various relationships or comorbidities between the ECG and ECHO parameters that contribute to HF; and finally, confirm those relationships via a Principal Component analysis (PCA), similarly to in.^27^ Understanding these findings together, determines more precisely the underlying physiological nature of abnormal functions, not just the symptoms of HF, for each phenotype, and associate them to novel bio-signals.

These key abnormalities appear to be addressed by a Breakthrough technology through the use of novel bio-signals and AI that emulates echocardiography findings^26^.

## 2 Analysis along HF categories

### 2.1 Background of HF categories

#### 2.1.1 HFpEF

HFpEF is widely recognized as a heterogeneous disorder, therefore the diagnosis relies on objective evidence of multiple structural and functional abnormalities in conjunction with LVEF≥50%: LVH, LAE, DD (elevated LV filling pressure), and elevated B-type natriuretic peptide (BNP) ^1^. Based on consensus recommendation from the Heart Failure Association (HFA) the initial HFpEF diagnosis relies on HF symptoms, standard ECG, standard ECHO, and a Natriuretic Peptides test that together comprises the *HFA-PEFF* algorithm ^4^.

LAE has been associated with abnormal DD,^28^ since DD contributes to left atrial remodeling ^29^. In the past, it was labeled “diastolic HF”, a suboptimal term given diastolic HF is not present in all HFpEF ^5^. Several complementary pathophysiologic mechanisms exist in HFpEF, including longitudinal LV systolic dysfunction (despite a normal LVEF), PH, abnormal ventricular-arterial coupling, abnormal exercise-induced vasodilation, extracardiac volume overload, and chronotropic incompetence.^5,30,16,17^ A PH relationship is confirmed by the *H*_*2*_*FPEF* score study, where the pulmonary artery systolic pressure >35mmHg was selected as one of the score elements.^9^ CAD, valve diseases, and PH are considered as primary comorbidities of HF,^16,31,32,17,15,33,34^ and the etiology of HFpEF relies on these abnormalities in the case of non-DD type HF.

HFpEF in a broader sense can be divided into various types, such as Coronary artery disease-HFpEF, Right heart failure-predominant HFpEF, Valvular HFpEF or AFib-predominant HFpEF (fully defined in ^5^). This wider definition of HFpEF is advantageous in primary care, since early detection of HF is as important as identifying the specific type of HF. However, the advanced diagnosis of HFpEF should be differentiated from the alternative causes of dyspnea such as HFrEF, valve disease, primary PH, or pericardial effusion ^4^.

#### 2.1.2 HFmrEF

HFmrEF is a recently separated HF class, thus fewer research results are available in the literature. Patients with HFmrEF have a different clinical profile, but one more similar to patients with HFpEF than with HFrEF ^35^; primarily mild LVSD, but with features of DD. The diagnostic criteria for HFmrEF include relevant structural heart disease, such as LVH or LAE, or DD beside symptoms and the mildly reduced EF (LVEF of 40-49%).^1^ HFmrEF is associated with different characteristics and a more favorable prognosis than HFrEF,^36^ has less comorbidity – as discovered by our analysis. HFmrEF patients who transitioned to HFrEF were more likely to have LAE including AFib and more comorbidities.^37^

#### 2.1.3 HFrEF

HFrEF is a complex clinical condition characterized by structural and/or functional impairment of the left ventricle, resulting in a decrease in heart pump function (LVEF≤40%) ^34^.

HFrEF is most commonly associated with DCM or CAD,^31^ LVSD and moderate AS commonly occur together; patients with moderate AS and concomitant LVSD are at high risk for clinical events including all-cause death, hospitalization for HF, and aortic valve replacement.^32^ Non-ECG detectable abnormalities such as severe valve regurgitations, including MR,^15^ AR,^33^ TR ^34^ increase the mortality of HF patients.

ECG in HFrEF patients is almost always abnormal, and most patients typically have a minimum of two but more typically three ECG abnormalities, with ECG LVH criteria being the most common abnormality.^38^ AFib and HF often occur together, and their combination is associated with increased morbidity and mortality compared with each disorder alone.^39^

#### 2.1.4 ALVSD

Although not an HF phenotype, mild Asymptomatic LVSD (ALVSD) is a predictor of adverse events mainly in subjects with combined DD.^40^ Patients with ALVSD (“Stage B” according to *ACC/AHA* of HF, NYHA CLASS I) have approximately half the mortality rate (5% annualized) of those with overt symptoms of HF, but their risk of death is 5 to 8 times higher than a normal age-matched population.^41,42^ Patients with ALVD are at high risk for developing HF, therefore, they should be considered potential HF patients.

In the Cardio-Phoenix database, the definitions of HF categories incorporate both LVSD and ALVSD.

### 2.2 Cardio-Phoenix database

The prevalence of HF and associated ECG and ECHO findings in primary care can be estimated from the Cardio-Phoenix database, because the collected medical data was specifically from a patient population at risk of Cardiovascular Disease (CVD) including HF. Patients are adults with an average age of 62.5, of which 51% are females, and have a minimum of three cardiovascular risks or an existing cardiac condition.

In accordance with the guidance,^3^ risks for HF (any type) include hypertension, cardiovascular disease, diabetes mellitus, obesity, known exposure to cardiotoxins, and family history of cardiomyopathy (see Stage A at-risk for HF in ^3,^). These risks were examined according to the criteria of patients entered into the database.

Databases consists of medical data from four clinical studies, completed at five European clinical centers, between 2011 and 2019. The database contains the bio-signals including ECG and ECHO images, measurements, and findings for each patient. The echocardiographic assessment was typically performed immediately or soon after bio-signal recording, but not more than a few days. There are some 24000 records with ∼30sec length signals. The ground truth of ECHO-findings (detailed in supplementary material) was established by cardiologist consensus and confirmed with ECHO measurements. The ECG measurements and findings are generated by state-of-the-art ECG algorithms based on a 12-lead ECG signal and confirmed by a minimum of one, but up to four, cardiologists.

In the Cardio-Phoenix database, the HF categories are defined based on the above-mentioned wider HF definitions, using all the available anamnesis/symptoms and clinical diagnoses supported by ECG- and ECHO-findings:

- HFpEF: symptomatic HF with LVEF>50% and DD or LAE or LVH presence, extended with symptomatic AF, symptomatic PH, symptomatic AS or MS, symptomatic CAD or WMA. This definition of HFpEF is consistent with the *TOPCAT, RELAX*, and *ARIC* studies.^43^
- HFmrEF: symptomatic HF with 40%<LVEF<50% extended with mild ALVSD confirmed with risk factors, HF scores, and co-morbidities of WMA or LAE, or LVH.
- HFrEF: symptomatic HF with LVEF<40% extended with ALVSD confirmed with risk factors, HF scores, and co-morbidities.

The overall database consists of 72.3% absent HF, 19.3% HFpEF, 5.9% HFmrEF and 2.5% HFrEF. The 72.3% absent category includes normal patients and patients with non-HF CVD – typically with a “mild” CVD condition. The overall HF patient is 27.7% in the database, and approximately half of the HF patient has HFpEF type (HFpEF 69%, HFmrEF 21%, and HFrEF 10%) similar to other studies.^6,43^

This categorization was the basis of the data analysis presented in Table 1 and sections (3, 4, and 5).

**Table 1.** Distribution statistics of ECHO and ECG interpretation in Cardio Phoenix Database along the four HF category – In each row, the worst or most abnormal (typically the maximum, but in some cases the minimum) values are highlighted with bold text, similarly, the highest standard deviation is also highlighted, which denotes the highest variability.

| Group | HF category |  | All |  | Absent HF |  | HFpEF |  | HFmrEF |  | HFrEF |  |
| --- | --- | --- | --- | --- | --- | --- | --- | --- | --- | --- | --- | --- |
|  | Field | Unit | M | STD | M | STD | M | STD | M | STD | M | STD |
| General and Body Size | Age | year | 62.6 | 13.8 | 59.7 | **13.8** | **72.0** | 9.4 | 67.9 | 10.3 | 67.2 | 12.1 |
|  | Female | % | 50.8 | 0.5 | 53.8 | 0.5 | 48.0 | 0.5 | 35.5 | 0.5 | **20.0** | 0.4 |
|  | BMI | kg/m<sup>2</sup> | 27.9 | 5.2 | 27.6 | 5.2 | 28.5 | 5.3 | 28.8 | 5.2 | **29.2** | **6.6** |
|  | BSA | m<sup>2</sup> | 1.9 | 0.2 | 1.9 | 0.2 | 1.9 | 0.2 | 1.9 | 0.2 | 2.0 | 0.3 |
|  | Height | cm | 167.5 | 9.1 | 167.7 | 9.2 | 166.2 | 8.7 | 167.7 | 8.7 | 169.4 | 9.9 |
|  | Weight | kg | 78.3 | 16.7 | 77.7 | 16.4 | 79.1 | 16.8 | 81.3 | 16.6 | **84.3** | **21.7** |
|  | Systolic BP | mmHg | 127.8 | 13.3 | 127.4 | 13.2 | **129.6** | 13.7 | 128.2 | 13.2 | 125.2 | **14.3** |
|  | Diastolic BP | mmHg | 78.6 | 9.9 | 78.8 | 9.6 | 77.8 | 9.7 | **78.9** | **12.9** | 77.4 | 11.6 |
| ECG measurements | HR | bpm | 70.0 | 13.6 | 68.4 | 11.8 | 73.7 | 16.8 | 76.3 | 17.7 | **77.8** | 15.8 |
|  | RR std | ms | 59.2 | 61.8 | 46.3 | 44.9 | 96.2 | 85.0 | 85.6 | 76.0 | **117.1** | 94.0 |
|  | LF/HF (0.15Hz) | ms | 1.6 | 0.9 | 1.8 | 0.9 | 1.3 | 0.8 | 1.2 | 0.8 | **1.1** | 0.6 |
|  | QRS axis | deg | 25.1 | 39.8 | 28.8 | 36.2 | 16.8 | 45.9 | 11.3 | 45.5 | **4.6** | **54.1** |
|  | PQ interval | ms | 181.8 | 37.7 | 175.2 | 28.6 | **201.7** | 51.1 | 195.7 | 52.4 | **204.7** | **52.7** |
|  | P interval | ms | 117.5 | 18.4 | 118.2 | 14.5 | 115.4 | 26.3 | 114.2 | 26.3 | **118.0** | **29.1** |
|  | P terminal force | mVms | 3.0 | 2.7 | 3.1 | 2.5 | 2.4 | 2.8 | 2.8 | 3.2 | **3.1** | **3.9** |
|  | QT interval | ms | 408.6 | 37.5 | 407.8 | 34.4 | **412.2** | 44.7 | 406.6 | **46.0** | 411.4 | 44.3 |
|  | QTc (Framingham) | ms+ | 425.1 | 26.0 | 422.1 | 23.3 | **433.0** | 30.1 | 432.3 | 31.1 | **440.7** | **33.9** |
|  | QRS interval | ms | 92.3 | 21.1 | 89.2 | 17.8 | 98.8 | 24.9 | 102.5 | 25.5 | **115.4** | **32.6** |
|  | VAT (Rwave peak time) | ms | 43.1 | 11.4 | 41.4 | 9.4 | 46.6 | 14.3 | 49.2 | 13.3 | **53.9** | 17.8 |
|  | Taxis | deg | 33.2 | 49.5 | 35.9 | 39.7 | 27.1 | 65.3 | **20.0** | 74.0 | 27.8 | **86.7** |
|  | Rsum (V1:V6) | uV | 4282 | 2138 | 4387 | 2091 | 4093 | **2248** | 3997 | 2165 | **3187** | 2204 |
|  | LVH Score | score | 166 | 201 | 128 | 159 | 256 | 242 | 320 | **293** | **353** | 264 |
| ECHO measurements | IVSd | mm | 10.2 | 1.8 | 10.0 | 1.6 | **11.2** | 2.0 | 10.4 | 1.9 | 10.0 | 2.1 |
|  | LVIDd | mm | 51.6 | 6.8 | 50.0 | 5.4 | 53.6 | 6.7 | 58.7 | 8.4 | **65.1** | 9.8 |
|  | LVIDs | mm | 33.8 | 6.8 | 31.9 | 4.7 | 35.3 | 5.8 | 44.4 | 6.4 | **53.1** | 8.7 |
|  | LVEF (Teichholz) | % | 63.3 | 8.7 | 65.5 | 6.5 | 62.5 | 6.8 | 47.6 | 2.4 | **37.5** | 4.6 |
|  | RWT | % | 34.4 | 6.7 | 34.7 | 6.3 | **35.5** | 7.3 | 31.4 | 7.3 | 27.2 | 8.4 |
|  | LVmass | g | 187.8 | 62.0 | 172.7 | 49.3 | 221.7 | 66.5 | 240.9 | 79.4 | **270.0** | **86.6** |
|  | LVMI (LVmass/BSA) | g/m<sup>2</sup> | 98.8 | 30.8 | 91.0 | 23.5 | 117.0 | 34.1 | 124.9 | 41.1 | **138.0** | 45.8 |
|  | LAV | mL | 36.6 | 20.8 | 29.9 | 11.1 | 51.9 | 25.0 | 58.4 | **33.7** | **74.5** | 31.4 |
|  | LAVI (LAV / BSA) | mL/m<sup>2</sup> | 19.3 | 11.1 | 15.8 | 5.6 | 27.4 | 13.2 | 30.7 | **19.4** | **38.5** | 18.5 |
|  | RAV | mL | 40.1 | 25.0 | 34.0 | 14.2 | 54.5 | 35.3 | 58.1 | 39.7 | **78.5** | **40.2** |
|  | RAVI (RAV / BSA) | mL/m<sup>2</sup> | 21.1 | 13.1 | 17.9 | 7.2 | 28.8 | 18.6 | 30.5 | **21.6** | **40.4** | 21.6 |
|  | AVpV | m/s | 1.4 | 0.5 | 1.3 | 0.4 | **1.7** | 0.8 | 1.5 | 0.8 | 1.4 | 0.6 |
|  | AVpGrad | mmHg | 8.9 | 9.4 | 7.4 | 5.6 | **14.3** | **15.6** | 11.4 | 14.4 | 9.0 | 10.2 |
|  | ARgrade | grade | 0.4 | 0.8 | 0.2 | 0.6 | **0.7** | 1.0 | 0.5 | 0.9 | **0.9** | 1.1 |
|  | ARjet Area | cm<sup>2</sup> | 0.6 | 1.8 | 0.4 | 1.3 | **1.3** | 2.7 | 0.9 | 1.9 | 1.4 | 2.7 |
|  | MVGE mean | mmHg | 2.0 | 1.4 | 1.8 | 1.1 | 2.5 | **1.9** | 2.5 | 1.7 | **2.6** | 1.7 |
|  | MV E Vmax | m/s | 0.8 | 0.3 | 0.8 | 0.2 | 0.9 | 0.3 | 0.9 | 0.3 | 0.9 | 0.3 |
|  | MV A Vmax | m/s | 0.8 | 0.3 | 0.8 | 0.2 | 0.8 | 0.4 | 0.7 | 0.4 | **0.6** | 0.4 |
|  | MV E/A |  | 1.1 | 0.4 | 1.0 | 0.4 | 1.1 | 0.6 | 1.2 | 0.5 | **1.5** | **0.8** |
|  | MRgrade | grade | 1.4 | 1.1 | 1.1 | 1.0 | 2.0 | 1.0 | 2.1 | **1.2** | **2.5** | 1.1 |
|  | MRjet/LAA | % | 13.5 | 15.1 | 10.6 | 12.7 | 20.0 | 16.7 | 23.5 | **21.5** | **28.1** | 18.5 |
|  | TRgrade | grade | 1.2 | 1.0 | 0.9 | 0.9 | **1.8** | 1.1 | 1.6 | 1.2 | **2.0** | 1.2 |
|  | TRjet/RAA | % | 12.5 | 16.1 | 9.5 | 13.8 | 21.1 | **19.2** | 18.6 | 17.9 | **24.1** | 18.0 |
|  | RVSP (mPAP) | mmHg | 18.9 | 11.8 | 16.3 | 8.3 | **26.5** | 15.8 | 24.8 | 16.6 | **29.5** | **18.1** |
|  | PEEF (maximum) | mm | 0.3 | 1.7 | 0.2 | 1.5 | 0.5 | 2.2 | 0.3 | 1.6 | **0.8** | **2.3** |
|  | RVOTend-distal | mm | 30.4 | 16.2 | 28.2 | 13.4 | 35.3 | 19.8 | 38.3 | 21.9 | **43.4** | 25.6 |
|  | RVOTprox | mm | 26.8 | 5.8 | 25.9 | 5.5 | 29.0 | 5.4 | 29.2 | 6.2 | **30.4** | 7.2 |
|  | RVDD M-mode | mm | 29.2 | 5.1 | 28.3 | 4.7 | 31.3 | 5.2 | 32.0 | 5.9 | **33.6** | 6.8 |
|  | RVDs M-mode | mm | 14.3 | 4.9 | 13.5 | 4.2 | 16.3 | 5.6 | 16.6 | 6.6 | **17.9** | 7.7 |
|  | RAD (A4C) | mm | 38.4 | 7.5 | 36.7 | 6.1 | 42.6 | 8.4 | 43.0 | 9.6 | 48.4 | 9.8 |
|  | RADI (RAD / BSA) | mm/m<sup>2</sup> | 20.4 | 4.2 | 19.5 | 3.4 | 22.6 | 5.0 | 22.5 | 5.7 | 24.9 | 6.0 |
|  | WM-score | score | 0.3 | 0.7 | 0.2 | 0.5 | 0.5 | 1.1 | 0.7 | 1.0 | 0.9 | 1.3 |
| HF Score | H<sub>2</sub>FPEF Score | score | 2.43 | 1.39 | 1.93 | 0.82 | 3.94 | 1.63 | 3.49 | 1.67 | 3.93 | 1.73 |
|  | HFA-PEFF score | score | 1.62 | 0.89 | 1.26 | 0.54 | 2.61 | 0.84 | 2.55 | 0.92 | 3.16 | 0.95 |
| ECHO-Finding | LVH |  | 14.0% |  | 7.8% |  | 34.6% |  | 22.9% |  | 33.6% |  |
|  | DCM |  | 9.9% |  | 3.7% |  | 15.2% |  | 44.3% |  | 76.0% |  |
|  | RVE |  | 4.6% |  | 2.2% |  | 8.9% |  | 12.5% |  | 25.3% |  |
|  | LAE |  | 8.8% |  | 0.5% |  | 29.7% |  | 30.6% |  | 54.3% |  |
|  | RAE |  | 6.5% |  | 1.2% |  | 17.7% |  | 24.2% |  | 42.8% |  |
|  | WMA |  | 4.5% |  | 0.6% |  | 4.5% |  | 38.3% |  | 44.1% |  |
|  | LVSD |  | 2.6% |  | 0.0% |  | 0.0% |  | 0.9% |  | 100.0% |  |
|  | DD Impaired Relaxation |  | 22.7% |  | 20.2% |  | 35.1% |  | 19.6% |  | 15.8% |  |
|  | DD Pseudo-normal |  | 3.5% |  | 2.0% |  | 6.5% |  | 9.2% |  | 11.5% |  |
|  | DD Restrictive Filling |  | 2.9% |  | 1.1% |  | 7.1% |  | 6.9% |  | 15.8% |  |
|  | AS |  | 5.7% |  | 1.8% |  | 20.1% |  | 10.5% |  | 7.9% |  |
|  | MS |  | 2.8% |  | 1.1% |  | 8.4% |  | 7.9% |  | 4.6% |  |
|  | AR |  | 7.0% |  | 4.2% |  | 15.4% |  | 10.5% |  | 24.0% |  |
|  | MR |  | 12.7% |  | 6.6% |  | 27.3% |  | 31.2% |  | 48.4% |  |
|  | TR |  | 9.9% |  | 4.6% |  | 24.1% |  | 22.0% |  | 36.8% |  |
|  | PH |  | 3.7% |  | 0.3% |  | 12.5% |  | 12.7% |  | 21.1% |  |
|  | **Abnormal ECHO** |  | 35.1% |  | 17.7% |  | 81.3% |  | 93.1% |  | 100.0% |  |
| ECG-Finding | S. Bradycardia |  | 3.2% |  | 3.5% |  | 2.9% |  | 1.7% |  | 1.6% |  |
|  | S. Tachycardia |  | 1.3% |  | 1.3% |  | 1.2% |  | 1.9% |  | 2.6% |  |
|  | AFib |  | 8.2% |  | 0.0% |  | 33.6% |  | 23.9% |  | 33.6% |  |
|  | PVC |  | 15.3% |  | 10.4% |  | 26.5% |  | 30.6% |  | 45.7% |  |
|  | PAC |  | 14.8% |  | 12.9% |  | 19.8% |  | 21.2% |  | 20.4% |  |
|  | Prolonged PR |  | 3.2% |  | 3.0% |  | 3.6% |  | 3.3% |  | 6.3% |  |
|  | Prolonged QT |  | 0.8% |  | 0.6% |  | 1.8% |  | 1.7% |  | 0.0% |  |
|  | BBB |  | 7.7% |  | 5.5% |  | 13.1% |  | 13.8% |  | 21.1% |  |
|  | ST-T deviation |  | 17.9% |  | 10.4% |  | 34.9% |  | 45.5% |  | 53.0% |  |
|  | Ischemic ST-T |  | 25.2% |  | 17.5% |  | 43.6% |  | 50.8% |  | 64.5% |  |
|  | MI criteria |  | 15.7% |  | 11.8% |  | 24.3% |  | 28.1% |  | 39.5% |  |
|  | LVH criteria |  | 9.1% |  | 5.1% |  | 18.6% |  | 23.9% |  | 27.0% |  |
|  | RVH criteria |  | 0.4% |  | 0.1% |  | 1.1% |  | 1.0% |  | 1.6% |  |
|  | Leftward Axis |  | 9.6% |  | 6.6% |  | 16.5% |  | 19.1% |  | 29.3% |  |
|  | Rightward Axis |  | 1.7% |  | 1.4% |  | 2.3% |  | 2.0% |  | 4.3% |  |
|  | **Abnormal ECG** |  | 40.9% |  | 28.6% |  | 72.4% |  | 78.8% |  | 92.8% |  |
|  | Borderline ECG |  | 25.0% |  | 28.4% |  | 16.4% |  | 16.3% |  | 5.3% |  |

### 2.2 Study Limitation

The right side of the heart is less represented by the included ECHO-parameters and by ECG-parameters even less, as such their place in the statistical results is limited.

Nonetheless, right-side heart failure and its structural, functional abnormalities and related pulmonary disease co-morbidities (Chronic obstructive pulmonary disease COPD) has attracted growing attention in the last few years. Right-heart side study through a bio-signal approach shows strong potential.

## 3 Prevalence statistics along four HF categories

### 3.1 Method

This work has been carried out in accordance with the Code of Ethics of the World Medical Association for experiments involving human subjects. Data used in the research were collection in a prior clinical study, already published with the Declaration of Helsinki state.^49^ To analyze prevalence along four HF categories a statistical analysis was performed on patient groupings into a) All – all the patient records, b) Absent HF records, c) HFpEF records, d) HFmrEF records and e) HFrEF records.

### 3.2 Results

Table 1 lists all the relevant information, measurements, and findings in the examined patient population.

#### 3.2.1 Highest prevalence

The listed measurements and findings show most of the abnormal mean values for the HFrEF category. Interestingly, the exceptions help us to better understand the HFpEF and HFmrEF categories:

- HFpEF shows the highest or significantly high value of Systolic Blood Pressure BP (it confirms other research, which shows HFpEF is prevalent in hypertensive populations^51^.), Interventricular Septal Diameter (IVSd), Relative Wall Thickness (RWT%), Aortic Valve Peak Velocity (AoV Vmax), AR Grade and Jet Area, Right Ventricular Systolic Pressure (RVSP), PQ and QT intervals, in addition, LVH, DDIM, AS, MS, AFib and Prolonged QT from findings. In addition, HFpEF shows the highest average age.
- HFmrEF shows the highest value of Diastolic BP, lowest T-wave axis, highest prevalence of Premature Atrial Contraction (PAC). In addition, significantly high prevalence of WMA, ST-T deviation, Myocardial Infarction (MI), and MS, which are closer to HFrEF values compared to HFpEF.
- Because of high variability and comorbidity rate, the HFrEF has the worst mean values and highest standard deviation for all the parameters, except for the parameters above mentioned. The distribution of several parameters confirms an earlier study result.^37^

#### 3.2.2 Characteristics of HFmrEF as a middle category

Until recently, the abnormalities that typically occur in HFmrEF compared to HFpEF and HFrEF have not been as comprehensively investigated.

The presented analysis reveals that HFpEF do not so much differ from HFmrEF in prevalence and disease severity as by disease type. HFpEF mostly includes DD, LVH, and mild/moderate valve disease patients, whereas HFmrEF trends towards patients with ischemic problems: WMA, MI, and ST-T deviation. Our analysis provides some relationships as presented in these statistics:

- LVH is represented within HFpEF and HFrEF but unexpectedly is a less typical indicator of HFmrEF
- DCM prevalence significantly increases in cases of HFmrEF from HFpEF and becomes frequent in HFrEF
- WMA is an important source of mildly reduced systolic function, with 8 times higher prevalence compared to patients with HFpEF. The additional prevalence increase is small compared to HFrEF.
- MI is typical in HFpEF, but even more so in patients with HFmrEF, where the prevalence is almost as high as in HFrEF, confirming a previous study.^37^
- PAC is more typical in HFmrEF and Premature Ventricular Contraction (PVC) for HFrEF.

#### 3.2.3 Typical abnormalities of three category

HFpEF group of patients are typically older, having hypertension, and obesity, where the typical structural abnormality is the LVH, the typical function abnormality is the DD, the typical hemodynamical problem is any aortic/mitral valve disease or PH, and the typical electrophysiological problem is AFib.

HFmrEF group of patients have similar comorbidities as HFpEF, but different types of problems: their typical structural abnormality is mild DCM, and the typical functional abnormality is WMA with either MI or ST-T deviation. The typical electrophysiological problems are AFib and PAC. Aortic valve disease is less typical compared to HFpEF, but Mitral/Tricuspid/Pulmonic are similar.

HFrEF group of patients have the highest levels of comorbidity and severity: the typical structural abnormalities are the DCM, left and right atrial and ventricular enlargement, the typical functional abnormality is LVSD, and the typical hemodynamical problem is Mitral/Tricuspid regurgitation and PH, and most common ECG abnormalities shows higher prevalence including the ECG LVH and ECG RVH criteria. The exception are sinus bradycardia, PAC, and prolonged QT.

Right-side heart abnormalities (RAE, RVE, RVH) show a significant increase in HFrEF categories. The overall comorbidity ratio shows an increasing trend with the decreasing of systolic function and exacerbation of HF. However, RV dysfunction is highly prevalent in HFpEF as well,^44^ moreover the recent COVID-19-related literature is increasingly referring to right side dysfunction with HFpEF.^45,46^

#### 3.2.4 The summary of HF co-morbidities prevalence

Table 2 shows the HF categories and typical co-morbidities in a summary analysis. The list of typical abnormalities or co-morbidities is listed using both the guideline and statistical analysis based on the presented data.

**Table 2.** HF category and related ECHO and ECG-based co-morbidities and abnormalities.

|  |  | Primary criteria<br>according to ESC Guideline, 2021 [1] |  | Secondary criteria<br>according to statistical analysis |  |
| --- | --- | --- | --- | --- | --- |
| HF<br>category | Symptoms<br>± Signs<br>criteria | LVEF<br>criteria | Structural /<br>Functional<br>criteria | wider definition in database -<br>applied for categorization | Typical co-morbidities<br>("+" denotes the disease plus the<br>previous ones) |
| Absent | No | ≥50% | - | - | Non-significant or mild<br>abnormalities |
| HFpEF | Yes |  | LVH<br>and/or LAE<br>and/or DD | symptomatic AFib<br>symptomatic PH<br>symptomatic AS or MS<br>symptomatic CAD or WMA | Regional WMA<br>Valve diseases (AS, AR)<br>PH<br>Arrhythmia (AFib)<br>Ischemic ST-T<br>Leftward Axis |
| HFmrEF | Yes | 41-49% | Global Mild LVSD | mild ALVSD with LVEF<50%,<br>but confirmed with risk factors,<br>HF scores and co-morbidities of<br>WMA or LAE or LVH | + Mild Dilated LV<br>+ Global WMA<br>+ More severe ischemia (CAD) |
| HFrEF | Yes | ≤40% | Global LVSD | ALVSD with LVEF<40%,<br>but confirmed with risk factors,<br>HF scores and co-morbidities | + Dilated LV (DCM)<br>+ More severe valve diseases<br>+ Right Heart Enlargement |
| Color Legend: |  |  |  |  |  |
| Primarily ECHO finding |  |  |  |  |  |
| Primarily ECG finding |  |  |  |  |  |
| Mixed ECG/ECHO finding |  |  |  |  |  |

(Abbreviations: AFib, Atrial Fibrillation; ALVSD, Asymptomatic LVSD; AR, Aortic Regurgitation; AS, Aortic Stenosis; CAD, Coronary Artery Disease; DCM, Dilated Cardiomyopathy; DD, Diastolic Dysfunction; ECG, Electrocardiogram; ECHO, Echocardiogram; HF, Heart Failure; HFmrEF, Heart Failure with mildly reduced LVEF; HFpEF, Heart Failure with preserved LVEF; HFrEF, Heart Failure with reduced LVEF; LAE, Left Atrial Enlargement; LVEF, Left Ventricular Ejection Fraction; LVH, Left Ventricular Hypertrophy; LVSD, Left Ventricular Systolic Dysfunction; MS, Mitral Stenosis; PH, Pulmonary Hypertension; WMA, Wall Motion Abnormality)

#### 3.2.5 ECG versus ECHO summary

To more clearly understand the differences between ECG and ECHO in predicting HF, we compare the Summary ECG with the Summary ECHO. In the database, both the ECG and ECHO interpretations are extended with a summary finding, listed in Table 1:

- ECG summary classifies the ECG as normal, borderline or abnormal
- ECHO summary classifies the ECHO as Normal, Mild or Abnormal: both moderate or Severe, and where Mild means a minimum of one Mild finding, Abnormal means a minimum of one Moderate/Severe finding

The best estimation reveals the limitation of ECG for HF: the ECG summary (Abnormal ECG) covers only 72.4% of HFpEF, 78.8% of HFmrEF, and 92.8% of HFrEF (average 81%), whereas the ECHO summary (Abnormal ECHO) has higher percentages, 81.3% for HFpEF, 93.4% for HFmrEF and 100.0% for HFrEF (average 92%), shown in Table 1. In other words, ECHO is far more sensitive and less inconclusive for diagnosing HF compared to ECG.

## 4 Prevalence trend comorbidity statistics

This statistical analysis shows how the co-morbidity is continuously increasing in the function of deterioration in LV systolic function: transitions are demonstrated between the three HF categories. Etiology and comorbidity analysis helps to understand the phenotype of HF and confirms the diagnosis.^43^

### 4.1 Method

In the presented analysis, consistent with the guidelines,^1,^ LVEF is used as an independent variable to estimate the prevalence of ECG and ECHO abnormalities. LVEF is the key continual measurement that separates the three categories of HF, which is split into 9 sub-categories with center values here, see in Fig. 1. Patients were grouped along these sub-categories and the prevalence of ECHO and ECG findings were calculated.

**Figure 1.**
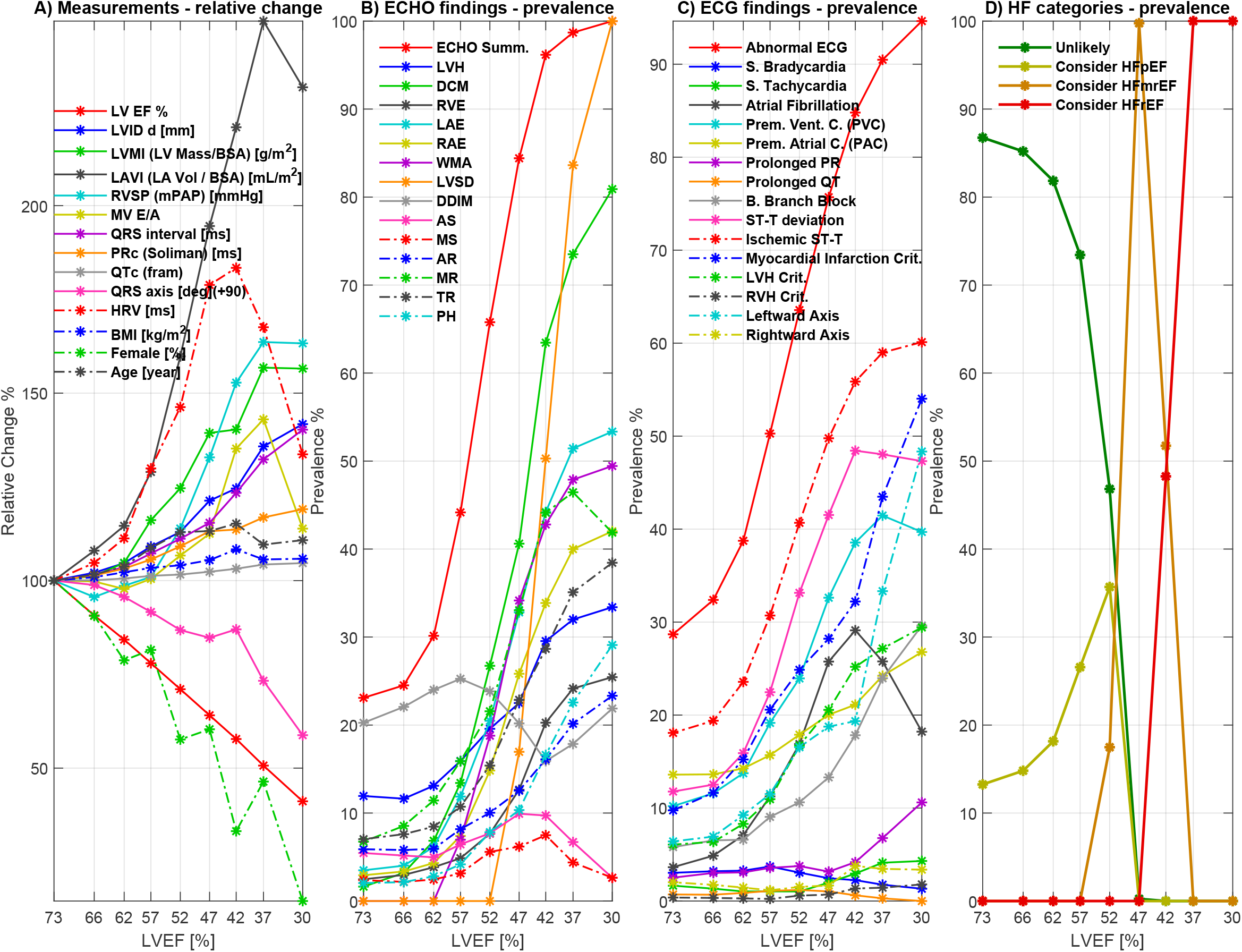
Comorbidity analysis in function of LVEF categories (in descending order): A) Average measurements: ECHO, ECG, BMI, Female, Age, B) ECHO-finding prevalence, C) ECG-finding prevalence, D) HF categories prevalence. (Abbreviations: AR, Aortic Regurgitation; AS, Aortic Stenosis; BMI, body mass index; DCM, Dilated Cardiomyopathy; DDIM, Impaired Relaxation of Diastolic Dysfunction; ECG, Electrocardiogram; ECHO, Echocardiogram; HF, Heart Failure; HFmrEF, Heart Failure with mildly reduced LVEF; HFpEF, Heart Failure with preserved LVEF; HFrEF, Heart Failure with reduced LVEF; HRV, Heart Rate Variability; LAE, Left Atrial Enlargement; LAVI, Left Atrial Volume Index; LVEF%, Left Ventricular Ejection Fraction (Teichholz); LVH, Left Ventricular Hypertrophy; LVIDd, End-diastolic Left Ventricular Diameter; LVMI, Left Ventricular Mass Index; LVSD, Left Ventricular Systolic Dysfunction; MR, Mitral Regurgitation; MS, Mitral Stenosis; MV E/A, E/A Wave Velocity; PH, Pulmonary Hypertension; QTc, Corrected QT interval; RAE, Right Atrial Enlargement; RVE, Right Ventricular Enlargement; RVH, Right Ventricular Hypertrophy; RVSP, Right Ventricular Systolic Pressure; TR, Tricuspid Regurgitation; WMA, Wall Motion Abnormality)

### 4.2 Results

The results are plotted in Fig. 1 together with the average value of some principal ECHO and ECG measurements:

- Graph A) shows a deteriorating trend of all the key ECG- and ECHO-measurements as a function of LVEF. Left Atrial Volume Index (LAVI) from ECHO and Heart Rate Variability (HRV) from ECG has the largest relative change, however HRV having maximum at LVEF=42%. Obesity, measured by average BMI, shows a slight increasing trend, and the percentage of Female patients drastically decreases with decreasing LVEF. This confirms that the HFpEF patient is biased to females, and the HFrEF patient biased to males. It is widely accepted that heart disease progresses with age, and the cardiac comorbidity statistics confirm both the disease progression and the increased comorbidity factor with increasing age.
- Graph B) shows constant increasing prevalence of almost all ECHO-findings, except: Impaired Relaxation (DDIM) having maximum around LVEF=57%; Aortic Stenosis (AS) and Mitral Stenosis (MS) having maximum around LVEF=42%. This confirms DDIM and aortic-mitral stenosis patients belong mostly to the HFpEF and HFmrEF categories.
- Graph C) shows constant increasing prevalence of almost all ECG-findings, except: S. Bradycardia has maximum around LVEF=57%; Sinus Tachycardia has minimum at LVEF=52%, ST-T deviation maximum around LVEF=42%; Rightward Axis having minimum at LVEF=57% and maximum at LVEF=42%. This confirms that typically, the HF patient has LVH with leftward axis, the Rightward Axis decreasing, but the typical right-side disease patients belong to HFrEF, that is why it is increasing again at the LVEF<45% groups.
- Graph D) shows the prevalence of each of the discussed HF categories, primarily separated based on LVEF. The maximum prevalence of HFpEF is around LVEF=52%, below 50% HFmrEF replaces the HFpEF classification and below 40% HFrEF replaces the HFmrEF classification.

## 5 PCA comorbidity statistics

### 5.1 Method

A Principal Component Analysis (PCA) was completed to reveal and unravel patterns of comorbidities, i.e. group of comorbidities associated with each other.^47^ PCA is a data reduction method that transforms the original set of variables into a smaller set of Principal Components (PCs), which are linear combinations of the original variables. PCs retain as much of the variability in the data set as possible, with the first component retaining the greatest amount of the variation present and the other components progressively retaining a decreasing amount of variation.^48^

In this PCA the largest variance is discovered in the data of the most important ECG, ECHO and patient body size measurements, which helps us to understand the interactions or co-morbidities of common cardiovascular diseases. Data was extended with two HFpEF scores: the *HFA-PEFF* score ^4,^ and the *H*_*2*_*FPEF* score ^9,^ in order to enable the determination whether the HF coincides with the direction of overall comorbidity rate.

Table 4 in supplementary material lists the 42 parameters included in PCA and the associated CVD.

### 5.2 Results

Fig. 3 shows the scatter plot of HF categories in function of *PCA1* and *PCA2*: the left graph A.) shows the four HF categories, where the centers are marked with black squares and numbers; the right graph B) shows the body size categories according to CHART Body Size Index (CBSI), where the CBSI<2.2 represent the smaller body size, 2.2<CBSI<2.4 represent the middle body size, and CBSI>2.4 represent the bigger body size (for more details about why CBSI is used instead of BSI can be seen in supplementary material). *PCA1* parameter is a good indication for HF with significant difference between Absent HF and HFpEF, and between HFpEF and HFrEF. There is no significant difference in *PCA1* between the two mild HF categories, HFpEF and HFmrEF.

**Figure 2.**
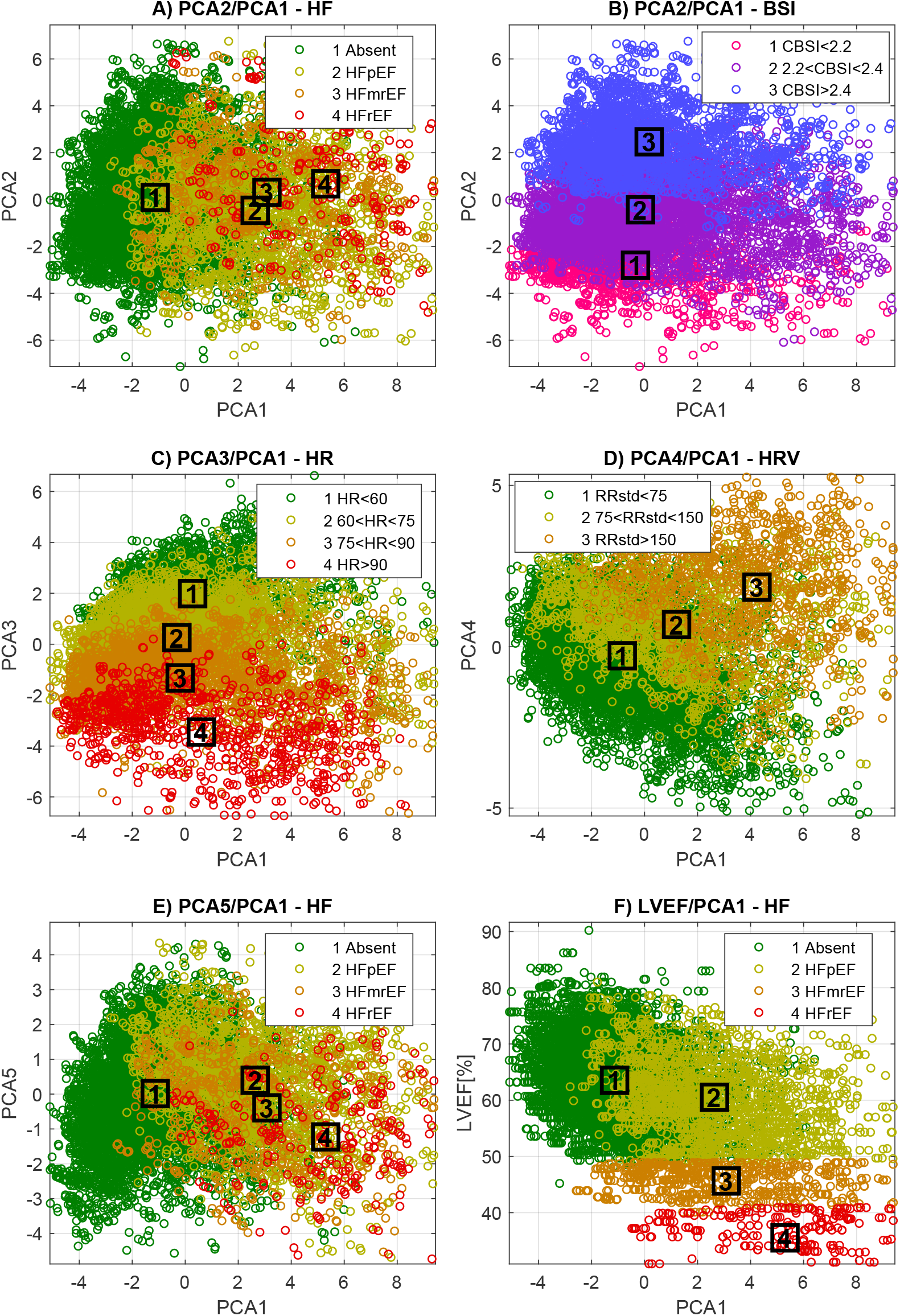
2D scatter plot of (A) HF categories in function of PCA1 and PCA2, (B) body size categories in function of PCA1 and PCA2, (C) HR categories in function PCA3 and PCA1, (D) HRV categories in function of PCA4 and PCA1, (E) HF categories in function of PCA5 and PCA1, (F) Four HF categories in function of LVEF and PCA1. (Abbreviations: BSI, body size index; CBSI – CHART body size index (similar to body surface area); HF, Heart Failure; HFmrEF, Heart Failure with mildly reduced LVEF; HFpEF, Heart Failure with preserved LVEF; HFrEF, Heart Failure with reduced LVEF; HR, Heart Rate; HRV, Heart Rate Variability; LVEF, Left Ventricular Ejection Fraction (Teichholz); PCA1, First Principal Component; PCA2, Second Principal Component; PCA3, Third Principal Component; PCA4, Fourth Principal Component; PCA5, Fifth Principal Component; RRstd, Standard Deviation of RR intervals)

**Figure 3.**
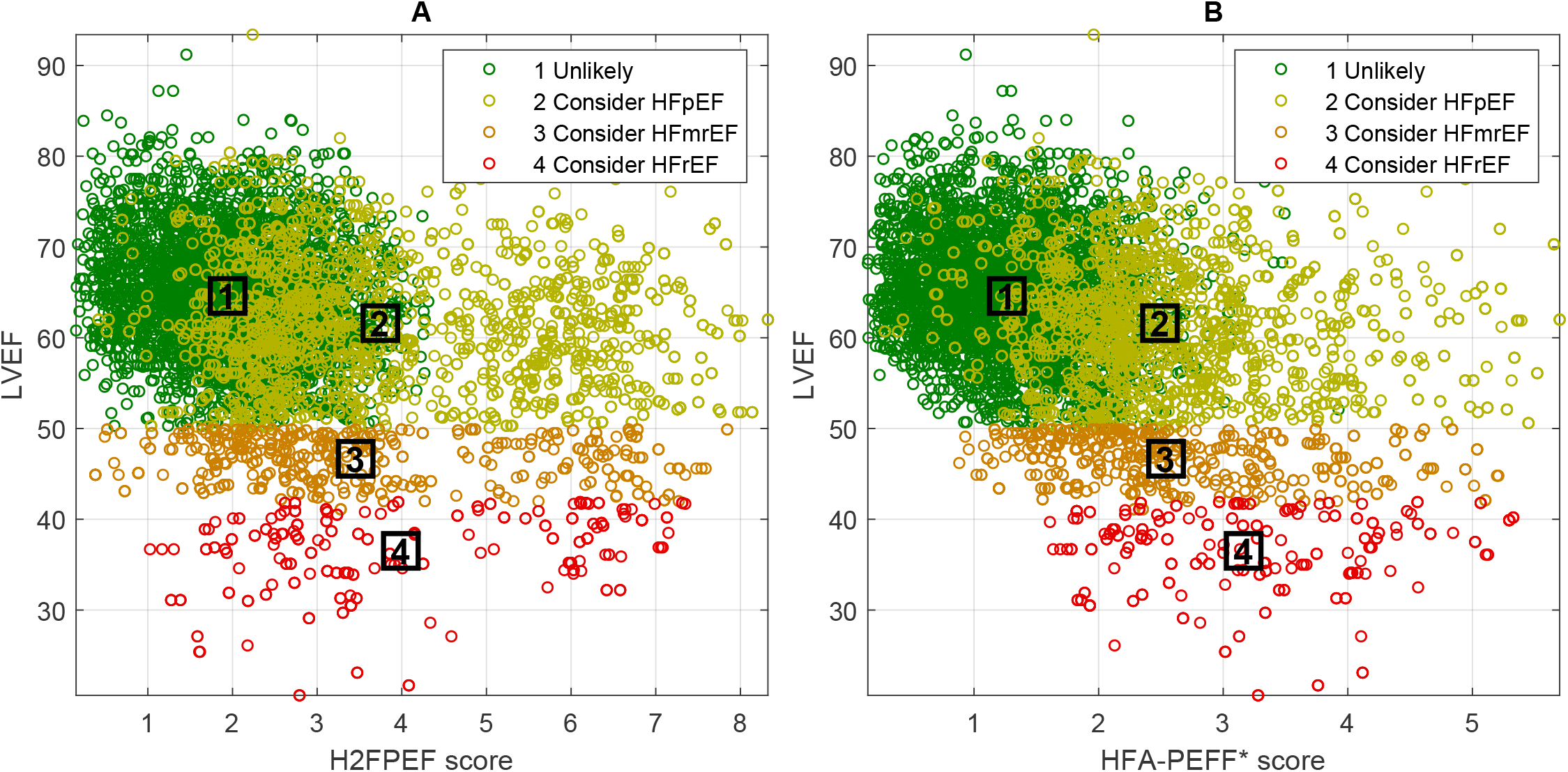
Scatter plot of HF categories in function LVEF (vertical) and HF scores (horizontal) (Abbreviations: HFmrEF, Heart Failure with mildly reduced LVEF; HFpEF, Heart Failure with preserved LVEF; HFrEF, Heart Failure with reduced LVEF; LVEF, Left Ventricular Ejection Fraction (Teichholz))

The second component *PCA2* represents the body size, which has not a pathological direction as *PCA1. PCA2* is at an independent perpendicular direction to *PCA1*, which carries the second biggest variance of data.

The scatter plot of the three heart rate (HR) categories - separated by average HR parameters - in function *PCA1* and *PCA3*, shown in graph C), and the Heart Rate Variability (HRV) categories – separated by standard deviation of RR intervals (RRstd) parameters in function of *PCA1* and *PCA4* (graph D.). The *PCA1* seems independent from HR but dependent from HRV. *PCA3* represents the HR value, and *PCA4* the HRV values. The scatter plot of four HF categories in function *PCA1* and *PCA5* shown in graph E). *PCA5* shows a DD related pathological direction, therefore is slightly higher separator between HFpEF and HFmrEF.

For more details about the presented PCs see Fig. 4 in supp. material, which shows the PCA coefficients of the first five components.

#### 5.2.1 Principal Components

Each of the PCs point in a specific direction, determined or represented by input parameters identified in Fig. 2. The presented first five PCA are considered the most important dimensions in the 42-dimension space. These first five PCA’s cover half of all the variance provided by the 42 dimensions: 18.2%, 11%, 9%, 5.6% and 4.8%, with the summary 49%.

Most of the common heart diseases have strong comorbidity, since all take part in the *PCA1*: HF and LVSD, atrial and ventricular enlargements, PH, MR, TR and LVH. The different natures of HFpEF and HFmrEF were not involved by the presented principal components. However, the LVEF can separate these two categories, see in graph F) in Fig 2., which shows the three types of HF in 2D space of *PCA1* and LVEF parameter.

The big role of *PCA1* can be observed in graph F), where *PCA1* distinguishes HFpEF from HF absent patients, which is not distinguishable by LVEF. LVEF can only distinguish HFmrEF and HFrEF from patients with LVEF>50%. Medical information incorporated in *PCA1* is required to make differential diagnosis of HFpEF.

The *PCA2*, 3 and 4 can be considered as independent components from *PCA1* and from each-other. These fulfill the supplementary directions: *PCA2* - body size, *PCA3* - HR and *PCA4* - arrhythmias.

The *PCA2* result confirms the important need of data normalization (for ECHO and ECG measurements) by body size or, at the very least, by gender. However, body size is best represented by BSA or CBSI because it relates more to the individual compared to the binary gender.

The *PCA5* is a supplementary diagnostic component for HF that helps to distinguish categories of DD and categories of HF. Compared to *PCA1, PCA5* shows higher distance between HFpEF and HFmrEF, as it represents the co-morbidity aspect of DD that is characteristic of HFpEF, despite the fact that in nearly half of the cases DD is absent ^5,^.

#### 5.2.2 ECG and ECHO factors of *PCA1*

The covariance pattern of ECG- and ECHO-measurements representing cardiac function was investigated by the PCA. The first principal component (*PCA1*) is interpreted as the overall comorbidity degree. The PCA established that the *PCA1* is representative of **structural abnormalities**: the atrial and ventricular enlargements. The direction of this component is mostly suitable to the HF severity categories: the HFpEF diagnostic scores are the most important participants in *PCA1*. This confirms the importance of ECHO over ECG in HF diagnoses, since these scores rely mainly on key ECHO-measurements: E/e’, LVMI, LAVI and RVSP.

The LAE defined by LAVI is the most representative ECHO-finding in the overall comorbidity rate, which coincides with the probability direction of HF. However, no single ECHO-measurement is suitable to identify all HF patients, hence the reason for the use of HF score approaches, where a set of relevant ECHO-findings are aggregated in order to account for the varying nature of HF. For example, ECG algorithms predicting LVEF<35%, like ^24,25^, target only HFrEF category, as such they are inherently insensitive for the larger group of HF patients found in primary care (HFmrEF and HFpEF).

In *PCA1*, the second group of coefficients are related to the functional abnormalities. Astonishingly, LVEF is the 14th input parameter according to the coefficient’s values, having similar impact as age, arrhythmia, prolonged PR, RVSP and Mitral/Tricuspid regurgitation. This means LVEF, age, arrhythmia, PR interval and valve diseases are less representative of the overall comorbidity compared to aggregated diagnostic scores and chamber enlargements.

## 6 Discussion

### 6.1 A novel Diagnosis, “Consider HF” for primary care

Unlike ECG, bio-signal derived ECHO-findings processed through a purpose designed AI system, can predict the structural and functional abnormalities and measurements, consistent with table 2 above, essential to the detection accuracy for HF,^49^ including low LVEF.

The real importance of bio-signal derived ECHO-findings lies in its simplicity as it makes detection of HF more immediate on patient presentation to Primary Care, precluding the use of purely symptomatic based clinical diagnosis that delays access to treatment. This means that on initial patient presentation to Primary Care, clinicians will be able to recognize HF, including classifying its severity and category, especially when symptoms remain inconclusive, confounding or ambiguous.

Cardio-HART™ system uses novel bio-signals analyzed by Artificial Intelligence, can predict 14 of the most prevalent and important ECHO-findings ^26,^ to support the novel diagnostic indications called “Consider HF”^1^. CHART is intended to: ***“detect potential abnormalities of a structural and functional nature related to a specific HF phenotype, independently from the current patient symptoms*.*”*** Where patient symptoms can be inconclusive, confounding and or ambiguous, Cardio-HART provides Echo finding based indications of cardiac abnormalities, including LVEF, consistent with this research such that a HF diagnosis should be considered.

The primary criteria of “consider HF” follows the LVEF categories as per the latest guideline ^1,^, while the secondary criteria confirm LVEF with the co-morbidities explored in this research and their risk factors including asymptomatic patients (ALVSD). Table 3 lists the supporting ECG and ECHO-findings provided by CHART to aid the “consider HF” determination.

**Table 3.** Supporting “Consider HF” categories by CHART.

| Reference categories of HF - specifically for indication in primary care | Primary criteria | Secondary criteria | CHART findings |
| --- | --- | --- | --- |
| “Consider HFpEF” | symptomatic HF with LVEF>50%, and DD or LAE or LVH | symptomatic AFib<br>symptomatic PH<br>symptomatic AS or MS<br>symptomatic CAD or WMA | AFib by ECG<br>PH by AI<br>AS, MS by AI<br>Ischemic ST-T by ECG, WMA by AI |
| “Consider HFmrEF” | symptomatic HF with LVEF<50% | mild ALVSD with LVEF<50%, but confirmed with risk factors, HF scores and co-morbidities of WMA or LAE or LVH | mild LVSD by AI<br>WMA by AI,<br>LAE by AI<br>LVH by AI |
| “Consider HFrEF” | symptomatic HF with LVEF<40% | ALVSD with LVEF<40%, but confirmed with risk factors, HF scores and co-morbidities | moderate LVSD by AI<br>DCM by AI<br>and all other ECG (BBB, ST-T) and AI-findings (RAE, RVE, MR, TR, AR) |

(Abbreviations: AFib, Atrial Fibrillation; AI, Artificial Intelligence, ALVSD, Asymptomatic LVSD; AR, Aortic Regurgitation; AS, Aortic Stenosis; BBB, Bundle Branch Block; CAD, Coronary Artery Disease; DCM, Dilated Cardiomyopathy; DD, Diastolic Dysfunction; ECG, Electrocardiogram; HF, Heart Failure; HFmrEF, Heart Failure with mildly reduced LVEF; HFpEF, Heart Failure with preserved LVEF; HFrEF, Heart Failure with reduced LVEF; LAE, Left Atrial Enlargement; LVEF, Left Ventricular Ejection Fraction; LVH, Left Ventricular Hypertrophy; LVSD, Left Ventricular Systolic Dysfunction; MR, Mitral Regurgitation; MS, Mitral Stenosis; PH, Pulmonary Hypertension; RAE, Right Atrial Enlargement; RVE, Right Ventricular Enlargement; TR, Tricuspid Regurgitation; WMA, Wall Motion Abnormality)

### 6.2 Verification by HFA algorithm

The accuracy of the “consider HF” categorization of HF is verified using the likelihood-based adaptations of HFA algorithm. The HF score-based algorithm relies mainly on echocardiographic measurements, and it is necessary because no single measurement is sufficiently decisive, but the HF probability can be well represented by aggregating the individual measurements ^4,9.^ Fig. 3 shows the three “consider HF” categories plus the “Absent HF” category in function of LVEF (vertical axis) and the two likelihood-based scores (horizontal axis): A) the *H2FPEF* Score; B) the *HFA-PEFF* Score. Both HF score adaptations are calculated using sigmoid probability function instead of strict binary threshold in order to ensure higher resolution and to better resolve borderline cases.

The two scores have similar results: The HFmrEF and HFrEF categories are primarily confirmed by LVEF, but the two HF scores also show non-normal ranges, similarly for HFpEF categories. The score results demonstrate the need of a second and independent component from LVEF, which can separate the HFpEF from the Absent HF category. This HFpEF component can be represented either by of the mentioned scores, but only in cases where ECHO is available, or, in cases where ECHO is not available, by the bio-signal predicted ECG/ECHO structural and functional parameters, as produced by CHART.

### 6.3 Conclusions

The prevalence-based comorbidity analysis shows that the discussed ECHO-findings are strong indicators for HF and its category. More precisely, for HF, ECHO shows **enlarged heart** with decreased myocardial contractility and general **abnormal heart functioning**.

Key set of ECHO-findings are sufficiently representative of HF when taken together as opposed to a single measurement, like LVEF. The two HF score techniques discussed in this study (*HFA-PEFF* ^4,^ and *H2FPEF* ^9,^) confirms this approach, as they rely on Echo-finding indicators for the critical components of their score, precisely to avoid reliance on LVEF. The use of LVEF only would lead to a limited HF prediction capability focusing only on the HFrEF category, ignoring HFpEF altogether and much of HFmrEF, resulting in a high incidence of false negative.

As Echocardiography is not widely or easily available in primary care, the key **structural** (chamber enlargements) and **functional** abnormalities related measurements are not available precluding the ability to diagnose HF other than through mainly symptomatic means. The standard ECG findings do not provide acceptable diagnostic power for HF, less so for its categorization.

As more patients with HF are expected to be living longer than ever before, healthcare priorities will increasingly shift to managing HF co-morbidities ^50^. A bio-signal based system, indicated for use in Primary Care, can address the lack of a suitable device. In such a bio-signal based approach the classifying of a set of ECHO-findings by AI is recommended, conformant with the 2021 guidelines set by the ESC for diagnosing of HF. In primary care the use of novel bio-signals of a physiological nature such as those found in Cardio-HART™ can provide a better understanding of the structural and functional abnormalities and thereby increase HF detection accuracy for all 3 types of HF, even in asymptomatic patients.

Further research is warranted to investigate in more detail the relationship between HF categories and right-side HF through the window of bio-signals, risk factors, detectable abnormalities, COPD and other co-morbidities and symptoms.

## Data Availability

The clinical data is for legitimate purposes upon request. Generally, however, the Official Study Report can be obtained upon request

## 7 Source of funding

The collection of the data used in this paper was previously collected in prior clinical studies funded in part by UVA Research Inc., no other funding was involved.

### 7.1 Authors Contributions

“G.B.Z. designed the analysis; E.SZ. and A.B.SZ. collected the data; G.K. validated the data; N.C. and D.T. contributed analysis tools; I.K. performed the analysis; A.Z. and M.A.N. wrote the paper.”

## Footnotes

1 © 2021 CardioPhoenix Inc., “consider HF” is also a trademark of Cardio-Phoenix Inc.

