## Supplementary Material for "Unraveling diagnostic co-morbidity makeup of each HF category as characteristically derived by ECG- and ECHO-findings"

### 1 Supplementary Material

#### 1.1 ECHO findings

Table 1 lists the ECHO-findings having cardiologist consensus ground truth, selected based on relevance for referral decision and high prevalence of diseases in the intended population (prevalence >3%).

**Table 1 – List of ECHO Findings having cardiologist consensus ground truth**

| ECHO finding | Baseline Criteria for MILD | Baseline Criteria for MODERATE/SEVERE |
|---|---|---|
| LVH | IVSd≥[10/11*]mm and LVMI>[100/115*]g/$m^2$ | IVSd≥[13/14*]mm and LVMI>[115/131*]g/$m^2$ |
| DCM | LVIDd>[53/59*]mm | LVIDd>[56/63*]mm |
| RVE | RVOTprox>30mm | RVOTprox>36mm |
| LAE | LAVI>30ml/$m^2$ | LAVI>40ml/$m^2$ |
| RAE | RAVI>30ml/$m^2$ | RAVI>40ml/$m^2$ |
| WMA | Mild Hypokinesis (WMscore≥1) | Hypokinesis, Akinesis, Dyskinesis (WMscore≥2 and LVEF<[54/52*]%) |
| LVSD | LVEF<[54/52*]% | LVEF<40% |
| DDIM | E/A<0.85 | E/A<0.70 |
| AS | AVpV>2.0m/s | AVpV>3.0m/s |
| MS | MVA<3.0$cm^2$ (MVGEm>3.7mmHg) | MVA<1.5$cm^2$ (MVGEm>5.0mmHg) |
| AR | ARgrade≥1 | ARgrade≥2 |
| MR | MRgrade≥1, MRjet/LAA>20% | MRgrade≥2, MRjet/LAA>30% |
| TR | TRgrade≥1, TRjet/RAA>20% | TRgrade≥2, TRjet/RAA>30% |
| PH | RVSP>40mmHg | RVSP>50mmHg |

* Different threshold for Female and Male [F/M], according to recommendations for cardiac chamber quantification by echocardiography

#### 1.2 PCA

Table 4 lists the parameters included in PCA and the associated CVD.

**Table 4 – List of PCA included measurements**

| Category | Abbr. | Unit | Parameter Description |
|---|---|---|---|
| ECHO parameters | LVEF | % | LV Ejection Fraction (Quinones Equation) |
| | LVMI | $g/m^2$ | Left Ventricular Mass Index |
| | LVIDd | mm | End-diastolic Left Ventricular Diameter (internal) |
| | RVOTprox | mm | Right Ventricular Outflow Tract Proximal |
| | LAVI | $mL/m^2$ | Left Atrial Volume Index |
| | RAVI | $mL/m^2$ | Right Atrial Volume Index |
| | WMscore | score | Wall Motion Score (1-Mild Hypokinesis, 2-Diffuse Hypokinesis, 3-Akinesis, 4-Dyskinesis, 5-Aneurism) |
| | E/A | | E/A Wave Velocity |
| | AVpV | m/s | Aortic Valve Peak Velocity (Vmax) |
| | MVGEm | mmHg | Mitral Valve Mean Gradient E wave |
| | ARgrade | grade | Rate of Aortic Regurgitation |
| | MRjet/LAA | % | Mitral Regurgitation Jet Ratio in Left Atrium Area |
| | TRjet/RAA | % | Tricuspid Regurgitation Jet Ratio in Right Atrium Area |
| | RVSP | mmHg | Right Ventricular Systolic Pressure |
| ECG parameters | HR-Mean | bpm | Heart Rate Mean Value |
| | RRmedian | s | median RR interval |
| | RR std | ms | Standard Dev. of RR intervals - SDNN |
| | PNN50% | % | Rate of \|dRR intervals\| > 50ms - PNN50 |
| | LF/HF | ms/ms | Low (0.04Hz-0.15Hz) per high (0.15Hz-0.4Hz) frequency parts of RR interval spectrum |
| | QRSaxis | deg/100 | QRS axis in frontal plane |
| | PQint | ms | PQ Interval |
| | PRa | ms | corrected PR interval (PRa=PR+a*(60/RR-70); Age<60:a=0.26, Age>60:a=0.42) |
| | Pint | ms | P Wave Interval |
| | Paxis | deg | P wave axis |
| | P-area | mVms | P negative area in V1 lead (P terminal force) |
| | QTint | ms | QT Interval |
| | QTc-Fram | ms+ | Corrected QT int. Framingham's: QTc=QT+0.154(1-RR) |
| | QTc-CHART | ms+ | Corrected QT interval, where the correction was made by age and body size |
| | QRSint | ms | QRS Interval |
| | VAT | ms | Global Rwave peak time (intrinsicoid deflection) |
| | STint | ms | ST interval |
| | Taxis | deg | T wave axis |
| | LVHsc | score | LVH modified Romhilt-Estes score |
| HF Scores | HFA-PEFFsc | score | HFA-PEFF diagnostic algorithm and scoring system for HFpEF (0-6) |
| | H2FPEFsc | score | H2FPEF score by Yogesh N.V. Reddy, developed to HFpEF (0-9) |
| Body Size | Age | year | Age |
| | Gender | | Gender (1 - Male, 2 – Female) |
| | BMI | $kg/m^2$ | Body Mass Index (weight[kg]/height[m]$^2$) |
| | BSA | $m^2$ | Body Surface Area (sqrt(Height[cm]*Weight[kg])/60) |
| | Height | cm | Height |
| | Weight | kg | Weight |
| | CBSI | m+ 1/5kg$^{0.27}$ | CHART Body Size Index (CBSI=Height[m]+0.2Weight[kg]$^{0.27}$) Note: CBSI is more appropriate body size index compared to BSI |

Fig. 4 shows the PCA coefficients of the first five components, where the input parameters are sorted by the absolute values of PCA1's coefficients.

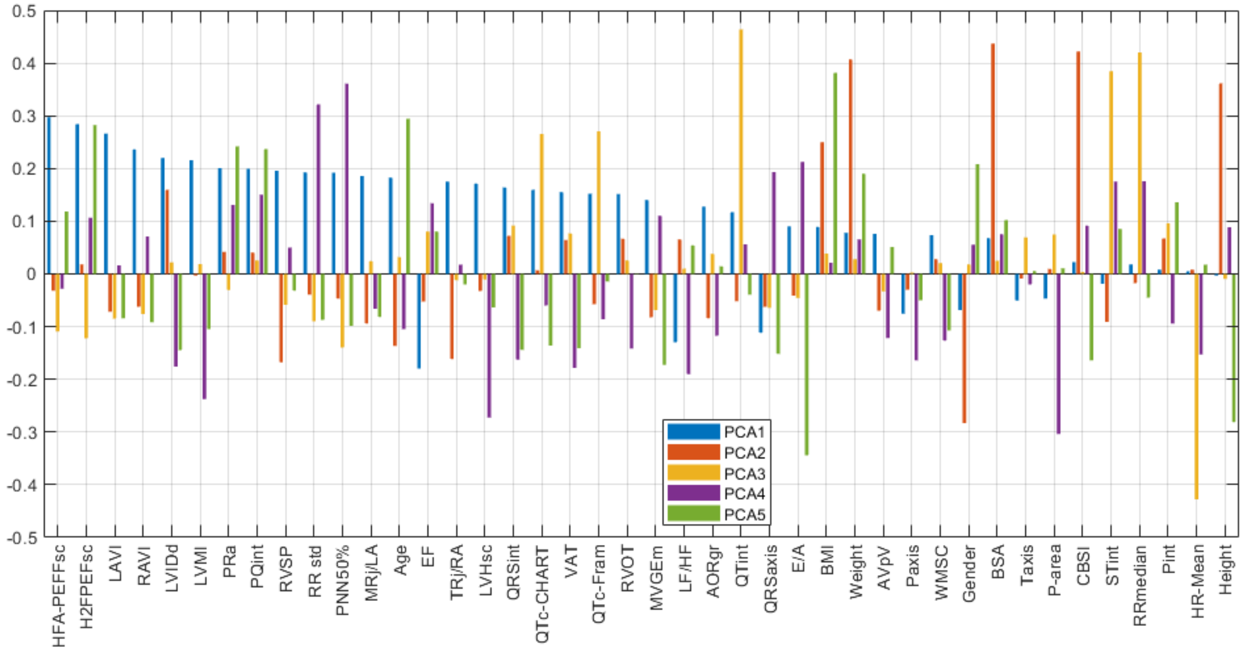


**Figure 1 – Linear Coefficients of PCA1, PCA2, PCA3, PCA4 and PCA5, calculated on the relevant ECHO, ECG, HFpEF scores and body size parameters**

#### 1.3 CHART Body Size Index – CBSI

Body Size Index (BSI) is required to be able to deal with body size differences across different sub-groups to minimize the risk of safety and effectiveness coming from demographic differences in interpretation of bio-signals or echocardiographic measurements.

The data presented in the EchoNormal study[1] that normalizing certain echo parameters with appropriate body size indexes, resulted in demographic differences (race and gender) that are practically negligible, or clinically meaningless differences.

Using the data provided in the EchoNormal study and the assumption that there should be a better indicator than Body Surface Area (BSA), this analysis resulted in the following Body Size Index named CBSI (**C**HART **B**ody **S**ize **I**ndex).

$$CBSI = heigth[m] + 0.2 \cdot weigth^{0.27}[kg]$$

The body size index currently used with echo devices, the BSA, still results in ~20% differences in normal values for the size measurements of left ventricle, which is significant. The CBSI lowers this difference to less than 4%, which represents a clinically

[1] http://www.echonormal.org/

meaningless difference resulting in a negligible change in risk leading to negligible changes in safety or effectiveness or to the benefit-risk profile of the demographic sub-groups.
